# Post-kala-azar dermal leishmaniasis (PKDL) drug efficacy study landscape: a systematic scoping review of clinical trials and observational studies to assess the feasibility of establishing an individual participant-level data (IPD) platform

**DOI:** 10.1101/2023.09.06.23295006

**Authors:** Sauman Singh-Phulgenda, Rishikesh Kumar, Prabin Dahal, Abdalla Munir, Sumayyah Rashan, Rutuja Chhajed, Caitlin Naylor, Brittany J. Maguire, Niyamat Ali Siddiqui, Eli Harriss, Manju Rahi, Fabiana Alves, Shyam Sundar, Kasia Stepniewska, Ahmed Musa, Philippe J Guerin, Krishna Pandey

## Abstract

**Background:** Post-kala-azar dermal leishmaniasis (PKDL) is a dermatosis which can occur after successful treatment of visceral leishmaniasis (VL) and is a public health problem in VL endemic areas. We conducted a systematic scoping review to assess the characteristics of published PKDL clinical studies, understand the scope of research and explore the feasibility and value of developing a PKDL individual patient data (IPD) platform.

**Methods:** A systematic review of published literature was conducted to identify PKDL clinical studies by searching the following databases: PubMed, Scopus, Ovid Embase, Web of Science Core Collection, WHO Global Index Medicus, PASCAL, Clinicaltrials.gov, Ovid Global Health, Cochrane Database and CENTRAL, and the WHO International Clinical Trials Registry Platform. Only prospective studies in humans with PKDL diagnosis, treatment, and follow-up measurements between January 1973 and March 2023 were included. Extracted data includes variables on patient characteristics, treatment regimens, diagnostic methods, geographical locations, efficacy endpoints, adverse events and statistical methodology.

**Results:** A total of 3,418 records were screened, of which 56 unique studies (n=2,486 patients) were included in this review. Out of the 56 studies, 36 (64.3%) were from India (1983-2022), 12 (21.4%) from Sudan (1992-2021), 6 (10.7%) were from Bangladesh (1991-2019), and 2 (3.6%) from Nepal (2001-2007). Five (8.9%) studies were published between 1981-1990 (n=193 patients), 10 (17.9%) between 1991-2000 (n=230 patients), 10 (17.9%) between 2001-2010 (n=198 patients), and 31 (55.4%) from 2011 onwards (n=1,865 patients). Eight (14.3%) were randomised clinical trials, and 48 (85.7%) were non-randomised studies. The median post-treatment follow-up duration was 365 days (range: 90-540 days) in 8 RCTs and 360 days (range: 28-2,373 days) in 48 non-randomised studies. Disease diagnosis was based on clinical criterion in 3 (5.4%) studies, a mixture of clinical and parasitological methods in 47 (83.9%) and was unclear in 6 (10.7%) studies. Major drugs used for treatment were miltefosine (n=636 patients), liposomal amphotericin B (L-AmB) (n=508 patients), and antinomy regimens (n=454 patients). Ten other drug regimens were tested in 270 patients with less than 60 patients per regimen.

**Conclusions:** Our review identified studies with very limited sample size for the three major drugs (miltefosine, L-AmB, and pentavalent antimony), while the number of patients combined across studies suggest that the IPD platform would be valuable. With the support of relevant stakeholders, the global PKDL community and sufficient financing, a PKDL IPD platform can be realised. This will allow for exploration of different aspects of treatment safety and efficacy, which can potentially guide future healthcare decisions and clinical practices.

**PROSPERO:** CRD42021295848

**Author summary:** Post-kala-azar dermal leishmaniasis (PKDL) is a dermatosis which mostly manifests after successful treatment of visceral leishmaniasis (VL) and is characterised by macular, papular, nodular, erythematous, or polymorphic rashes. PKDL is a public health problem in VL endemic areas, as recent infectivity studies show that *L. donovani* parasites can be found in PKDL lesions and remain infectious to sandfly vectors. There are numerous gaps in our existing knowledge of PKDL, including its pathology, immunology, and risk factors associated with therapeutic outcomes. Currently recommended treatments are either expensive (liposomal amphotericin-B), have raised safety concerns (especially for antimony regimens), or require long treatment duration (e.g. miltefosine). In order to scope the measure of evidence supporting therapeutic efficacy recommendations for PKDL patients, we conducted a systematic literature review. Our systematic review identified 56 PKDL studies describing 2,486 patients, with a majority of the studies (31 studies and 1,865 patients) published from 2010 onwards. The Infectious Diseases Data Observatory (IDDO) already have an established data platform for VL, and the <u>IDDO VL data platform</u> currently hosts a critical mass of data from efficacy trials in VL conducted over the past 20 years. Based on the identified volume of data, with a substantial number of studies being relatively recent, we believe that the establishment of a PKDL data platform is feasible. Creating a platform to facilitate the sharing of the datasets would enable in-depth IPD meta-analyses with existing data to address several knowledge gaps of PKDL and guide future research priorities. With the help of relevant stakeholders, the global PKDL community and sufficient resources, a PKDL data platform can be realised and help address key research gaps.

## Introduction

Post-kala-azar dermal leishmaniasis (PKDL) is a skin disorder that mostly manifests among patients after successful treatment of visceral leishmaniasis (VL). The disease is characterised by macular, papular, nodular, erythematous, or polymorphic rashes [1-3], and its incidence is estimated to range from 5-10% of treated VL patients in the Indian sub-continent (ISC) to 50% in Sudan [2]. It may also develop in asymptomatic carriers of *Leishmania donovani* [4]. Furthermore, an estimated 10-23% of diagnosed PKDL cases have no previous history of VL [5].

Geographical differences in clinical and epidemiological characteristics of PKDL cases have been reported [1]. Maculopapular rashes are seen in 90% of the cases in East Africa (EA), whereas macular rash is observed in a large proportion of the cases in the ISC [6]. Most PKDL cases in Sudan self-cure, while treatment is only recommended for chronic (> 6 months) and severe cases [7, 8]. On the other hand, treatment is always required for a successful cure of PKDL cases in the ISC [1]. The recommended drug regimens for the treatment of PKDL in India are currently either miltefosine (MF) for 12 weeks or 60-80 doses of amphotericin B deoxycholate (amphotericin B) at 1 mg/kg body weight per day over four months [9]. In EA, treatment is initiated mainly in severe cases with either sodium stibogluconate (SSG) (20 mg/kg/day) for up to 2 months or a 20-day regimen of liposomal amphotericin B (LAmB) (2.5 mg/kg/day) [2, 10]. However, these treatments are far from ideal in terms of safety, efficacy, adherence, and duration [2, 11-13]. More recently, shorter regimens with LAmB used as monotherapy or in combination with other drugs have been evaluated [14, 15].

PKDL is a public health problem in VL endemic areas as the PKDL skin lesions contain the parasite and can be an infective reservoir [16, 17]. Therefore, prompt detection and treatment of PKDL cases are important for the control and elimination of the disease. Data generated from clinical trials and longitudinal studies serve as the backbone for evidence-based case management and help identify knowledge gaps to guide future research. Existing systematic reviews on efficacy studies of PKDL have so far focused on a limited set of therapeutic options [18], and to date, there has been no comprehensive systematic review to assess the landscape of prospective PKDL clinical studies aimed at describing drug efficacy. Poverty-related neglected tropical diseases such as VL and PKDL have historically received little research and development investment [15]. Consequently, these diseases which often impact people experiencing poverty are characterised by fewer studies, a smaller number of patients, and the small volume of generated data can remain in silos. The scarce data from completed PKDL studies remains an underutilised source of information that can help answer specific research priorities and identify knowledge gaps.

This systematic review aimed to systematically identify PKDL treatment efficacy studies in the literature and evaluate the feasibility of establishing a global PKDL individual patient data (IPD) platform. The specific objectives were:

a. To identify PKDL treatment efficacy studies (clinical trials and observational) and summarise the study characteristics.
b. To assess the technical feasibility of developing a global IPD database for PKDL, i.e., how much of the IPD are likely to be retrieved from the studies identified in this review.

## Material and methods

### Protocol and registration

The protocol for this scoping review was prospectively registered with PROSPERO (CRD42021295848)[20] and was conducted in accordance with the Preferred Reporting Items for Systematic-Reviews and Meta-Analyses Extension for Scoping Reviews (PRISMA-ScR) guidelines [19]. PRISMA-ScR checklist is attached as **supplemental file 1**.

### Information sources and search strategy

A systematic scoping review of published literature was first conducted on 2^nd^ June 2020 and updated in full on 3^rd^ March 2023 by an expert librarian (EH) to identify PKDL efficacy studies by searching the following databases: PubMed, Scopus, Ovid Embase, Web of Science Core Collection, WHO Global Index Medicus, PASCAL, Clinicaltrials.gov, Ovid Global Health, Cochrane Database, and CENTRAL, and the WHO International Clinical Trials Registry Platform (ICTRP). Only prospective human studies with PKDL diagnosis, treatment, and follow-up measurements between 1973 and March 2023 were included. The search was not restricted by language of publication. The details of the search strategy used and the time period of the search are presented in **supplemental file 2**.

### Eligibility criteria and study selection

Studies in this review were screened in two stages, namely title and abstract screening and full-text screening, according to pre-defined inclusion and exclusion criteria. All clinical trials and prospective observational studies, in which patients were treated for PKDL and followed-up, were eligible for inclusion. The inclusion of studies was not restricted by the age of patients, the drug regimen used as an intervention, the comparator used, the length of post-treatment follow-up, or treatment outcome to be able to include all relevant studies. The following formed part of the exclusion set: studies published as case-reports, case-series, retrospective studies, diagnostic or prognostic studies, cross-sectional studies, letters, and studies that included the wrong population (non-PKDL, non-human), were published before the year 1973, had duplicate records, and publications with full-texts not retrievable. See the PROSPERO registration (CRD42021295848) for complete eligibility criteria. All the articles were independently screened by two reviewers (AM, RK, SR, and SSP) in a blinded fashion using Covidence Systematic Review Software (RRID: SCR_016484; December 2022)[20]. Any disagreement was resolved through consensus with a third reviewer.

### Data extraction

Variable and data dictionaries with extraction rules were developed for constructing a REDCap (RRID:SCR_003445) electronic data capture tool [21, 22]. From each of the articles deemed eligible for inclusion, the following variables were extracted: study title, publication year, country, study design, patient characteristics (age, gender), inclusion and exclusion criteria, diagnostic methods, treatment regimens, follow-up, efficacy endpoints, adverse events, and statistical methodology. The detailed metadata for the database is provided in **supplemental file 3**. The REDCap extraction tool was piloted on 10% of eligible studies by two authors (CN, RK) to validate the database and extraction rules. Two authors (AM, RK) each extracted data from half of the eligible studies and cross-verified each other’s extraction. Extracted data from ten percent of the randomly selected article was verified by two additional authors (RC, SSP). Any discordances in data extraction were discussed for clarity and consistency with a third reviewer (PD, SSP) and resolved through consensus.

### Data analysis and reporting

Descriptive characteristics of the included studies were presented. Details of patient characteristics were reported, and the details of the treatment regimen administrated, including dosage and duration of treatment, were summarised. Descriptive analysis was carried out using R software (R Project for Statistical Computing; RRID: SCR_001905).

### Risk of bias assessment

The primary aim of this study was to systematically scope the literature on PKDL studies to gauge whether the establishment of a PKDL IPD platform is feasible. Meta-analysis of study results was not intended, and no specific clinical research questions were posed. Therefore, the results from this review are limited to a descriptive analysis of study and patient characteristics of identified literature, and risk of bias assessment is therefore not conducted.

## Results

### Search outcomes

The review identified a total of 3,418 records across the literature databases searched. After excluding 22 duplicate records, a total of 977 records were eligible for title and abstract screening. Of the latter, 514 records did not meet the eligibility criteria, leaving 441 records for full-text screening. After reviewing the full texts, 66 studies were considered eligible for inclusion in this review. Ten of the 66 studies are currently ongoing and remain unpublished, leading to a total of 56 unique studies describing 2,486 patients included in this review (**Figure 1**).

**Figure 1:**
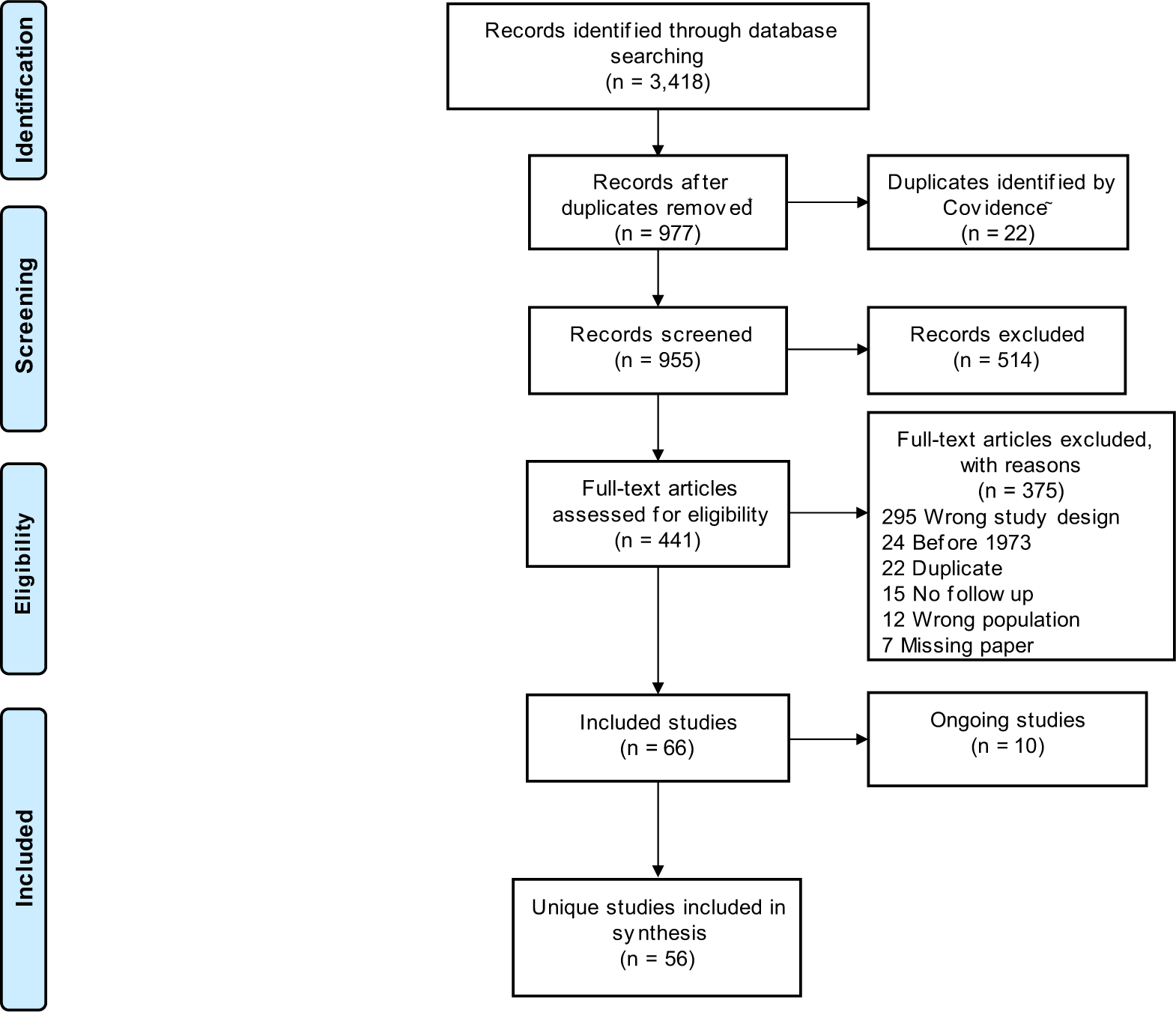
PRISMA flow-diagram. Legend: WHO ICTRP =World Health Organization International Clinical Trials Registry Platform. A separate search was carried out on WHO ICTRP as the server was unavailable during the time when other databases were searched.

### Publications by region and time-period

Of the 56 included studies, 36 (64.3%) were from India (1983-2022), 12 (21.4%) from Sudan (1992-2021), 6 (10.7%) were from Bangladesh (1991-2019), and 2 (3.6%) from Nepal (2001-2007) (**Figure 2**). Overall, these studies enrolled 2,486 patients, of which 1,512 (60.8%) were from India, 662 (26.6%) were from Bangladesh, 265 (10.7%) were from Sudan, and 47 (1.9%) were from Nepal. There were no studies identified between 1973-1980, 5 (8.9%) studies were published between 1981-1990 (n=193 patients), 10 (17.9%) between 1991-2000 (n=230 patients), 10 (17.9%) between 2001-2010 (n=198 patients), and 31 (55.4%) from 2011 onwards (n=1,865 patients) (**Figure 2**).

**Figure 2:**
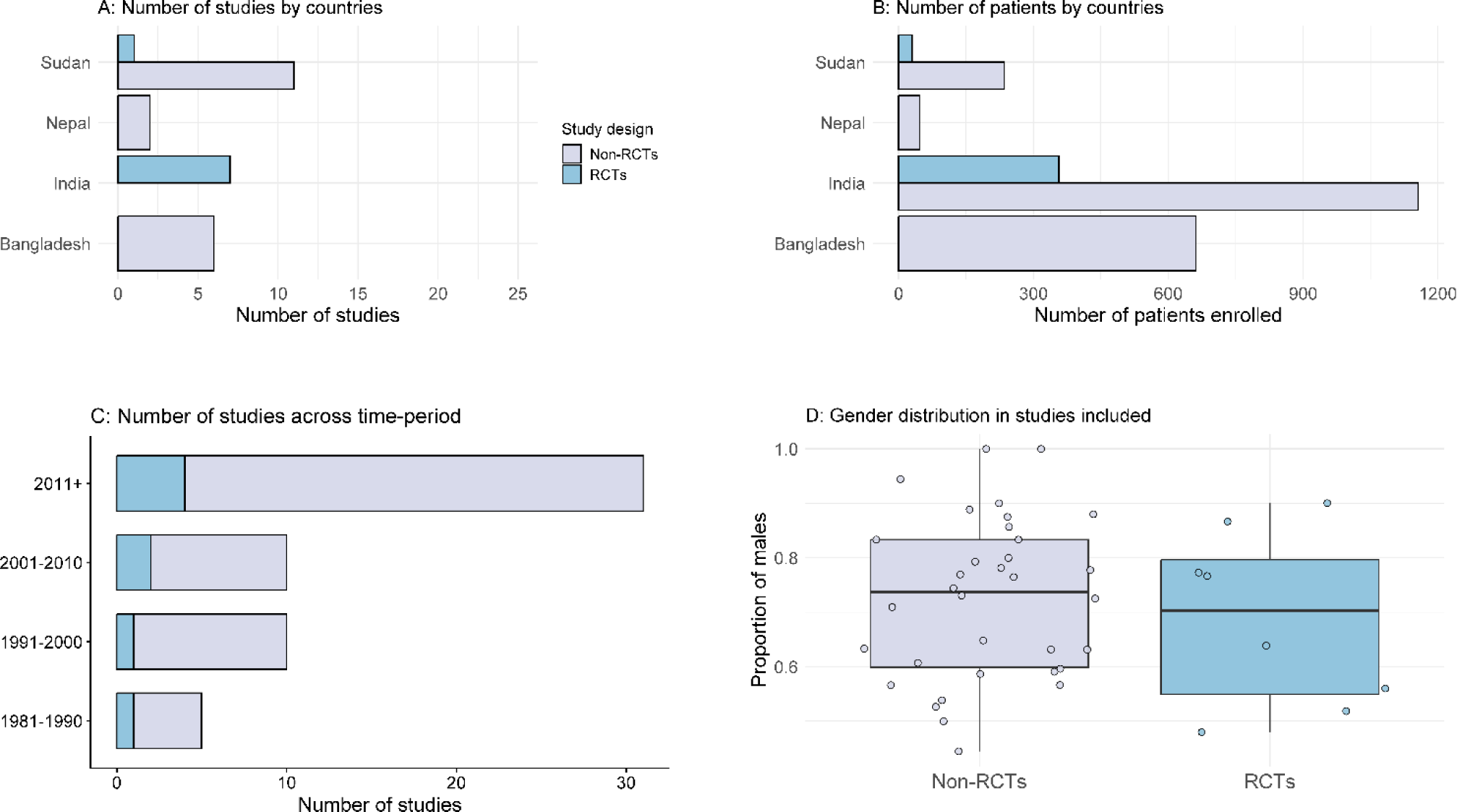
Characteristics of the studies included in the review.

### Study design

Of the 56 studies included, 8 (14.3%) were randomised studies (1987-2021) and the remaining 48 (85.7%) were non-randomised studies (**Figure 2**). Of the 8 RCTs (n=386 patients), 1 was an open label trial, 1 was single blinded and blinding was not stated in the remaining 6 trials. The median number of patients recruited per study across 8 RCTs and 48 non-randomised studies were 33 (range 10 - 108) and 26 (range 2 - 280), respectively. The median post-treatment follow-up duration was 365 days (range: 90-540 days) in 8 RCTs and 360 days (range: 28-2,373 days) in 48 non-randomised trials (**Table 2 and Table 3**).

### Patient characteristics

Overall, patients of all age ranges were enrolled in 28 (50.0%) studies, only adults (15+ years) were enrolled in 4 (7.1%) studies; and the age range of the patients included was not clear in the remaining 24 (42.9%) studies. The age-range of the patients included in the studies is depicted in **Figure 3**, for the 32 studies on which the age range data was presented. The gender distribution of the study population by study type is presented in **Figure 3**. Of the 2,486 patients enrolled, 1,270 were males, 650 were females and gender were not stated for the remaining 566 patients. The median proportion of males enrolled was 73.7% [range: 4.4%-100%] in 48 observational studies and 70.3% [range: 48.0% −90.0%] in 8 RCTs. None of the studies explicitly reported enrolling PKDL patients with HIV, pregnancy, para kala-azar, or any other comorbidities and it was unclear if they were excluded. Previous treatment for VL was clearly confirmed in 203 (52.6%) of the 386 patients enrolled in 8 RCTs and 936 (44.6%) of the 2,100 patients enrolled in 48 non-randomised studies.

**Figure 3:**
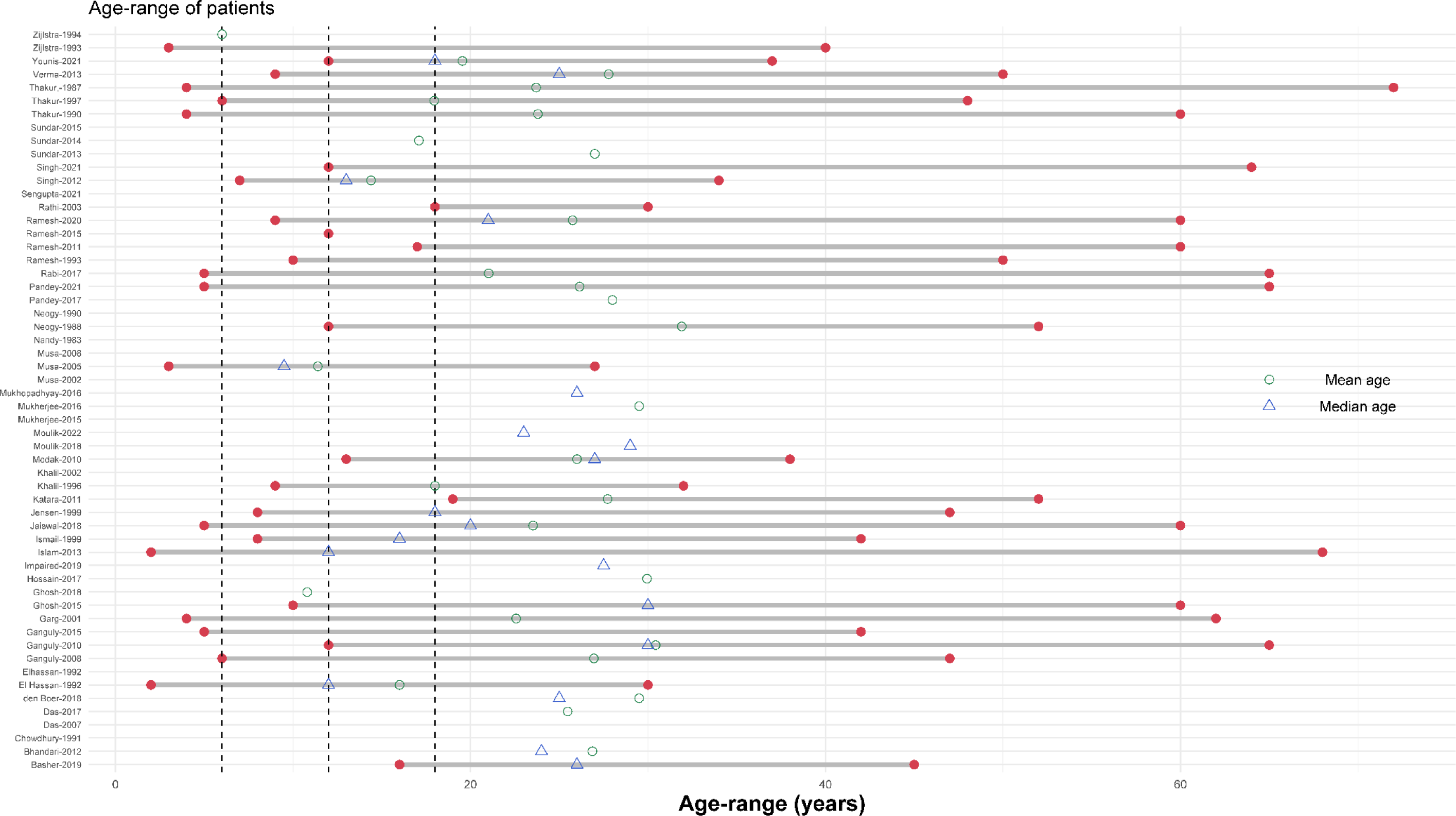
Age-range of the participants included in the review. Legend: The vertical dotted line presents age of 5, 10, and 15 years respectively. Each row represents a single study.

### Diagnostic methods

Only clinical criterion was adopted as a diagnostic method for disease identification in 3 (5.4%) nonrandomised studies, a mixture of clinical and parasitological demonstration in 48 (83.9%) studies on the whole, and the diagnostic method was unclear in 7 (10.7%) nonrandomised studies (**Table 1**). Overall, 43 (76.8%) studies used one or more laboratory tests for confirmed PKDL diagnosis in addition to clinical presentations (**Table 1**). All randomised clinical trials used one or more confirmatory tests for PKDL diagnosis. Microscopic demonstration of amastigotes in skin biopsies (slit or snip skin smears) in addition to other methods (clinical, serological, PCR, or culture) was used in 37 (66.1%) studies. Polymerase chain reaction (PCR) was used in 21 (37.5%) studies, while serological test was used in 27 (48.2%). A further breakdown of the diagnostic methods used over time-period is presented in **Figure 4**.

**Figure 4:**
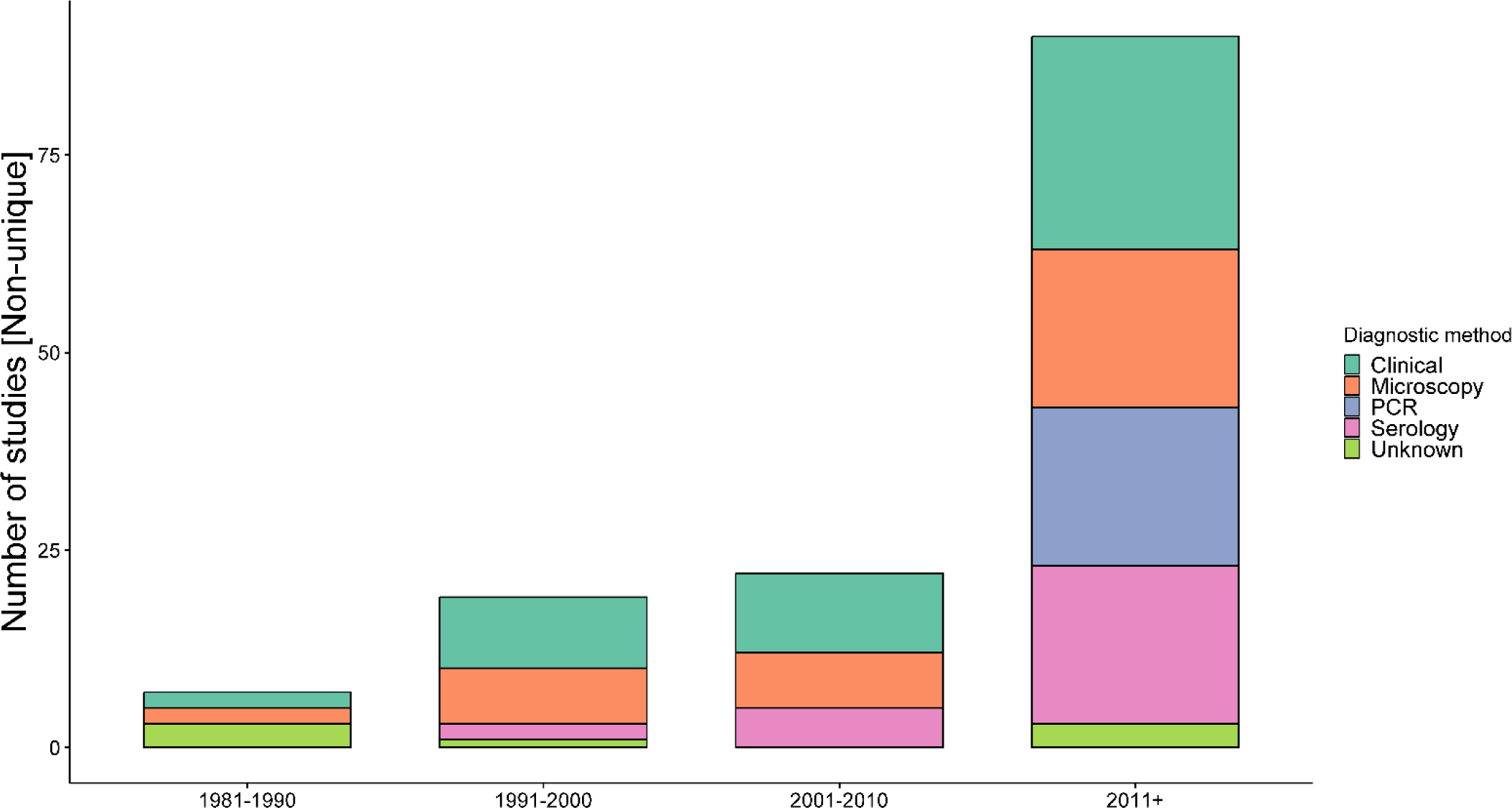
Diagnostics methods adopted over time. **Legend:** The number of studies is non-unique as a single study can contribute to multiple categories. See Table 1 for different combination of diagnostics methods adopted.

**Table 1:** Diagnostic methods used for PKDL across studies.

|  | **Randomised studies**<br>**(8 studies; 386 patients)** |  | **Non-randomised studies**<br>**(48 studies; 2,100 patients)** |  | **Overall**<br>**(56 studies; 2,486 patients)** |  |
| --- | --- | --- | --- | --- | --- | --- |
| **Diagnostic method for**<br>**PKDL confirmation** | Number of<br>studies (%) | Number of<br>patients (%) | Number of<br>studies (%) | Number of<br>patients (%) | Number of<br>studies (%) | Number of<br>patients (%) |
| Clinical + Microscopic | 4 (50%) | 184 (47.7%) | 8 (16.7%) | 169 (8%) | 12 (21.4%) | 353 (14.2%) |
| Clinical + Microscopic +Serological + PCR | 2 (25%) | 150 (38.9%) | 9 (18.8%) | 345 (16.4%) | 11 (19.6%) | 495 (19.9%) |
| Unknown | - | - | 7 (14.6%) | 116 (5.5%) | 7 (12.5%) | 116 (4.7%) |
| Clinical + Microscopic +Serological | - | - | 6 (12.5%) | 119 (5.7%) | 6 (10.7%) | 119 (4.8%) |
| Clinical + Serological | 1 (12.5%) | 22 (5.7%) | 5 (10.4%) | 441 (21%) | 6 (10.7%) | 471 (18.9%) |
| Clinical + Microscopic + PCR | - | - | 5 (10.4%) | 343 (21%) | 5 (8.9%) | 343 (13.8%) |
| Clinical | - | - | 3 (6.3%) | 125 (6%) | 3 (5.4%) | 125 (5.0%) |
| Clinical + Serological + PCR | - | - | 3 (6.3%) | 156 (7.4%) | 3 (5.4%) | 156 (6.3%) |
| Clinical + Microscopic + Culture | 1 (12.5%) | 30 (7.8%) | - | - | 1 (1.8%) | 22 (0.9%) |
| Clinical + PCR | - | - | 1 (2.1%) | 185 (8.8%) | 1 (1.8%) | 185 (7.4%) |
| Microscopic + Serological | - | - | 1 (2.1%) | 101 (4.8%) | 1 (1.8%) | 101 (4.1%) |

Column percentage presented in parenthesis; PCR= polymerase chain reaction

### Drug regimens

Thirteen different treatment regimens were adopted across 82 treatment arms. Details of treatment regimens at study arm levels were available for only 1,876 patients and it was missing for the remaining 610 patients. The most widely used drug regimen was miltefosine which was administered to 636 (33.9%, 636/1,876) patients followed by liposomal amphotericin B (L-AmB) (n=508/1,876, 27.1%), sodium stibogluconate (n=268/1,876, 14.3%), sodium antimony gluconate (n=186/1,876, 9.9%) and 10 other drug regimens, all them enrolling less than 60 patients (**Table 4**; **Figure 5**).

**Figure 5:**
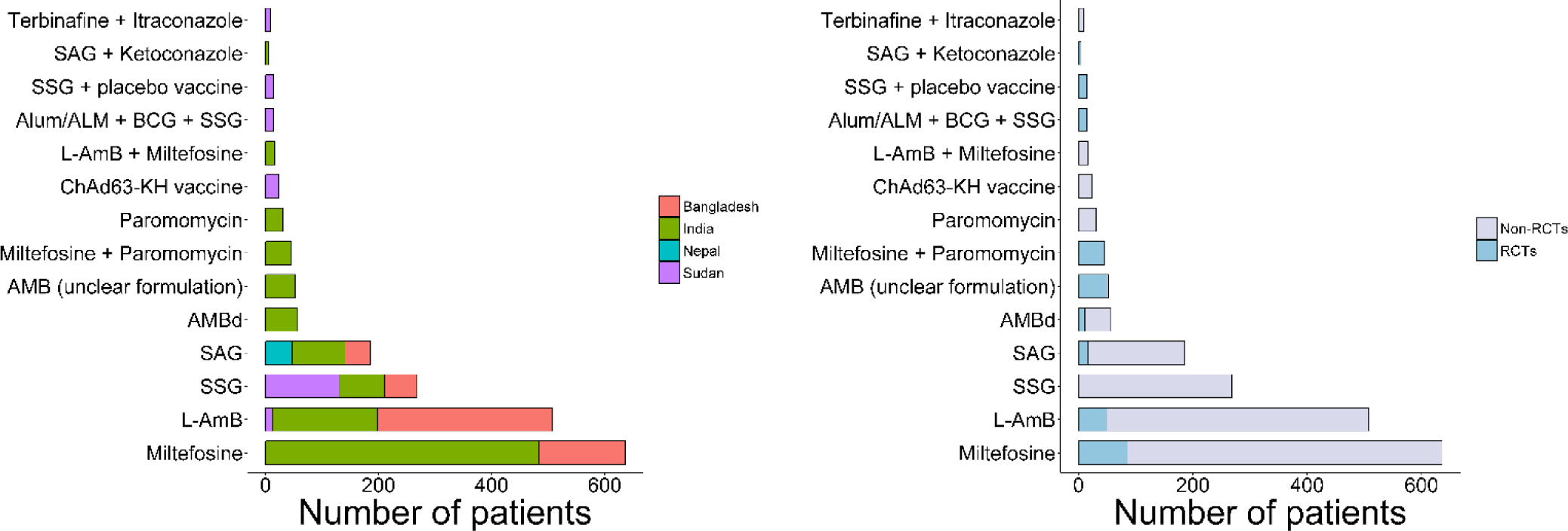
Drug regimen adopted by country and study design. SAG = sodium antimony gluconate; SSG=Sodium stibogluconate; L-AmB= Liposomal Amphotericin B; AMBd = Amphotericin B deoxycholate

Six different drugs were tested in three combination therapies: liposomal amphotericin B + miltefosine (n=16/1,876, 0.9%), terbinafine + itraconazole (n=9/1,876, 0.5%), and sodium antimony gluconate + ketoconazole (n=5/1,876, 0.3%). Vaccine as a therapy was tested among 54 (2.9%) PKDL patients in two studies from Sudan. ‘No treatment’ as a natural healing effect was used as a comparator arm, including eight (0.4%) PKDL patients in one study.

There was variation in daily mg/kg dose and overall treatment duration within the same regimen (**Figure 6**). L-AmB was tested at a total dose of 15 mg/kg over 3 weeks in 1 study (n=280 patients), 30 mg/kg over 3 weeks in 3 studies (n=118 patients) and a total dose of 50 mg/kg in 1 study (n=12 patients). Miltefosine was tested at 100 mg/day for 3 months in 4 studies (n=176 patients) with dosage ranging from 100-150 mg/day over 2-4 months period, while the daily target dose of antimony regimens ranged from 10-20 mg/kg/day over 1 to 4 months period (**Figure 6**).

**Figure 6:**
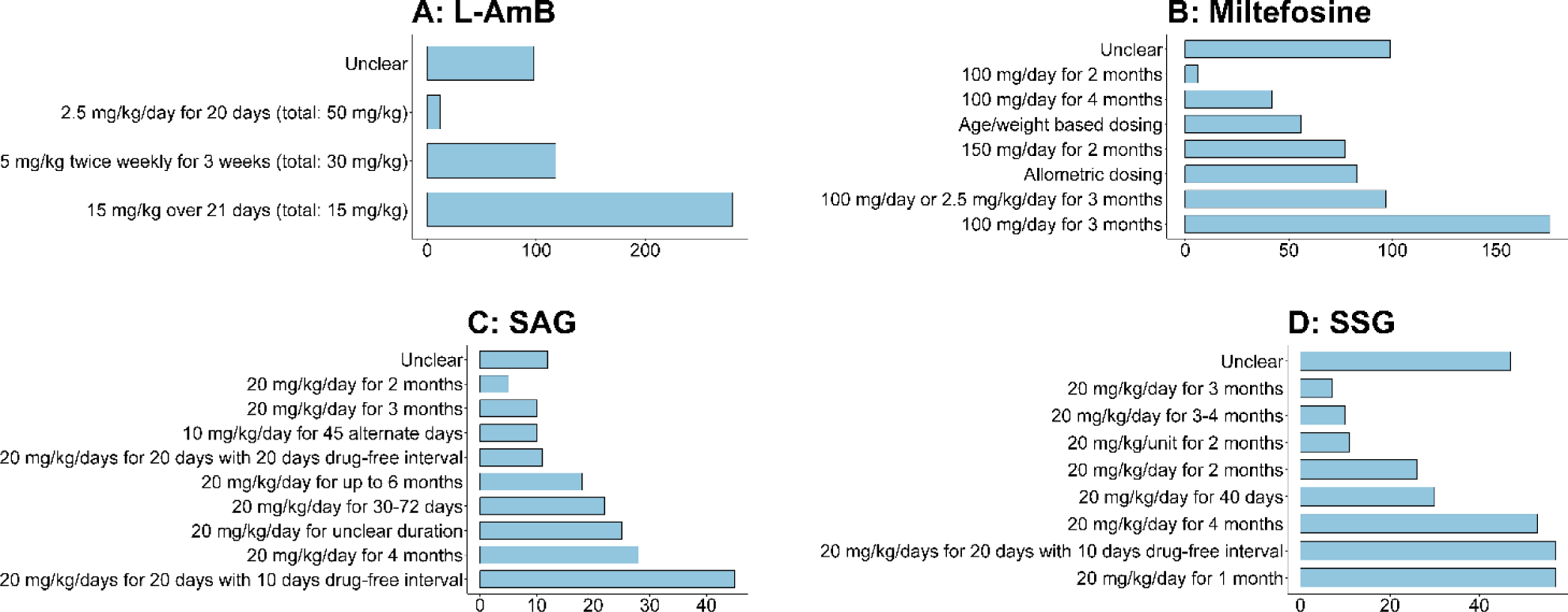
Number of patients treated with different total mg/kg dosage of pentavalent antimony, Miltefosine, and L-AmB. SAG = sodium antimony gluconate; SSG=Sodium stibogluconate; L-AmB= Liposomal Amphotericin B

### Assessment of cure in RCTs (n=8 studies)

Three RCTs clearly defined the initial cure criteria used; all these included disappearance/clearance or regression of lesions at the end of the therapy while the remaining 5 studies didn’t explicitly state the criterion for initial cure (**Table 2**). Final cure (or cure) was defined as complete disappearance of the lesions in 1 study, complete disappearance of the lesions along with parasite score of 0 in 4 studies, as absence of relapse in 1 study, and the definition was not clear in the remaining 2 studies (**Table 2**). Details regarding the proportion of patients who achieved initial and final cure are presented in Table 2, when reported.

**Table 2:** Details of randomised studies included in the review (8 studies; 386 patients)

| IDDO ID | Author-year | Country | Age-range (years) | Total enrolled | Drug regimen | Follow-up (days) | Criteria for Initial cure | Criteria for final cure | Initial cure estimates* | Final cure estimates* |
| --- | --- | --- | --- | --- | --- | --- | --- | --- | --- | --- |
| 1 | Thakur et al. 1997 [44] | India | 6-48 | 22 | Arm 1: Amphotericin B (i.v., 1 mg/kg/day on days 1-20, 21-40 and 61-80, separated by a 20-days interval)<br>Arm 2: SAG (i.m., 20 days courses at 20 mg/kg.day up to a daily dose of maximum of 850 mg), with a 20-days drug-free intervals) | 365 | Not stated | Complete clearance of lesions | Not reported | 100% (11/11)<br>63.6% (7/11) |
| 15 | Pandey et al. 2021 [45] | India | 5–65 | 100 | Arm 1: L-AmB (30 mg/kg, in six doses of 5 mg/kg, biweekly for 3 weeks)<br>Arm 2: Miltefosine (2.5 mg/kg or 100 mg/day for 12 weeks) | 365 | Disappearance of nodular and papular lesions and/or no appearance of new macular lesions together with grade 0 parasitaemia score in skin tissue after treatment | Complete disappearance of all papular/nodular lesions, complete or almost complete resolution of macular lesions, no reappearance of new lesions and grade 0 parasitaemia scores or insignificant parasitic burden in the skin at 12-month follow-up | Arm 1: 98% (49/50)<br>Arm 2: 100% (50/50) | Arm 1: 74.5% (35/47)<br>Arm 2: 86.9% (40/46) |
| 20 | Musa et al. 2008 [46] | Sudan | Not stated | 30 | Arm 1: Four weekly doses of 100 ug Alum/ALM + BCG (one-tenth of the 0.1 ml dose used for TB vaccination), i.m. | 90 | Not stated | Not stated | Not reported. | Arm 1: SSG+Alum/ALM+BC G, 87% (13/15) healed completely within |
|  |  |  |  |  | Arm 2:<br>Four doses of the vaccine diluent (placebo) |  |  |  |  | 60 days and the remaining 13% (2/15) showed considerable improvement. Arm 2: SSG + placebo, 53% (8/15) of the patients healed completely within 60 days, one patient (7%) showed considerable improvement. |
| 25 | Pandey et al. 2017 [47] | India | Not stated | 30 | Arm 1:<br>Intramuscular injection of paromomycin (11 mg/kg) daily for 10 days for two courses with a gap of 15 days. + Miltefosine capsule orally (2.5 mg/kg/day) daily for 10 days for two courses with a gap of 15 days.<br>Arm 2:<br>Intramuscular injection of paromomycin (11 mg/kg) daily for 10 days for three courses with a gap of 15 days. + Miltefosine capsule orally (2.5 mg/kg/day) daily for 10 days for three courses with a gap of 15 days. | 365 | Clearance of parasite from the dermal lesions at the end of treatment (EOT) | Disappearance of lesions and grade 0 parasite score at one year follow up | Arm 1: 100% (15/15)<br>Arm 2: 100% (15/15) | Presented at study level: 83.3% (25/30) |
| 38 | Sundar et al. 2013 [48] | India | Not stated | 36 | Arm 1:<br>Miltefosine was administered at a target dose of 2.5 mg/kg/day for 12 weeks (12-week group) utilising 50-mg capsules: two capsules for patients greater or equal to 25 kg or one capsule for <25 kg. | 365 | Not stated | Final efficacy responses were determined at 12 months after the end of treatment. Cure was defined as a clinical score = 0 for all three | Not reported | Arm 1: 93% (14/15)<br>Arm 2: 81% (13/16) |
|  |  |  |  |  | Arm 2:<br>Miltefosine was administered at a target dose of 2.5 mg/kg/day for 8 weeks (8-week group) utilising 50-mg capsules: two capsules for patients greater or equal to 25 kg or one capsule for <25 kg. |  |  | locations and a parasitological score = 0 when last measured after treatment. |  |  |
| 43 | Thakur et al. 1987 [49] | India | 4–72 | 108 | Sodium stibogluconate intramuscularly daily for 120 days:<br>Arm 1: 10 mg/kg body weight<br>Arm 2: 15 mg/kg<br>Arm 3: 20 mg/kg | 365 | Not stated | Patients were seen every month for 12 months after the end of treatment, and cure was assumed when there had been no relapse during follow up. | Eighteen patients in Arm 1 and 12 in Arm 2 were not cured at 120 days, so their dose was increased to 20 mg/kg; all were cured after a further three months. | Three patients required more than 120 days' treatment with the 20 mg dose; all were cured with further treatment, two with another 40 injections and one, with extensive lesions, with another 80 injections. Two patients in group C developed electrocardiographic changes, and the drug was stopped for 20 days; the changes were resolved and the drug was restarted. One patient developed arthralgia with a plasma urate concentration of 0.49 mmol/l; this responded to allopurinol and indomethacin while the treatment was continued. All patients tolerated the longer course of treatment. |
| 50 | Rathi et al. 2003 [50] | India | 18–30 | 10 | Arm 1: Sodium antimony gluconate (20 mg/kg bodyweight daily)<br>Arm 2: Sodium antimony gluconate (20 mg/kg per day) and ketoconazole orally (200 mg twice daily) | 540 | Not stated | Undefined | Not reported | Arm 1: the nodules and/or plaques showed approximate 80-85% clinical improvement, and macules showed 25-30% improvement.<br>Arm 2: 90-95% clinical improvement in the nodules and/or plaques and 25-30% in macules. |
| 59 | Das et al. 2017 [51] | India | 5–65 | 50 | Arm 1: Amphotericin B (0.5 mg/kg daily for 20 infusions for 3 courses at an interval of 15 days between each course)<br>Arm 2: Amphotericin B (1.0 mg/kg, alternate days for 20 infusions for 3 courses at an interval of 15 days between each course) | 365 | Initial cure as complete regression of papular and nodular lesions and/or no appearance of new macular lesions, together with a grade 0 parasite score in the dermal lesion and/or insignificant parasite burden by qPCR at the end of treatment. | Final cure constituted of disappearance of clinical signs and symptoms for PKDL, total re-pigmentation of macules and Grade 0 parasite score in dermal lesions at twelve months follow-up. | Arm 1: 100% (23/23)<br>Arm 2: 100% (22/22) | Arm 1: 95.7% (22/23)<br>Arm 2: 95.5% (21/22) |

i.v. = intravenous; i.m. = intramuscular; LAmB = Liposomal amphotericin B

### Assessment of cure in non-randomised studies (n=48 studies)

Of the 48 non-randomised studies, only 3 (6.3%) studies clearly defined initial cure (as clinical improvement by days 30 and/or 90 from treatment onset) and 19 (39.6%) studies clearly defined the criteria used for assessment of the final cure (**Table 3**). All 19 studies required complete or partial regression/healing of lesions for achieving cure with 5 studies explicitly requiring complete healing of lesions and 2 studies additionally requiring negative (or negligible) parasitaemia using PCR (**Table 3**).

**Table 3:** Details of the non-randomised studies included in this review (48 studies; 2,100 patients)

| IDDO ID | Author-year | Country | Age-range (years) | Total enrolled | Drug regimen | Follow-up period (days) | Definition of initial cure | Definition of final cure |
| --- | --- | --- | --- | --- | --- | --- | --- | --- |
| 2 | Nandy A et al. 1983 [52] | India | Unknown | 12 | Sodium antimony gluconate (age-based) | 180 | Undefined (clinical and immunological response measured but no specific details of clinical response presented) | Undefined (clinical and immunological response measured but no specific details of clinical response presented) |
| 3 | Musa AM et al. 2005 [12] | Sudan | 3 – 27 | 12 | LAmB (2.5mg/kg/day for 20 days) | 90 | ‘Clinical improvement’ (reduction in the severity and type of rash and/or darkening of any hypopigmented lesions by day 30 and/or day 90). | ‘Clinical cure’ (complete healing of the skin lesions and re-pigmentation of any previously hypopigmented lesions by day 30 and/or day 90). |
| 4 | Moulik S et al. 2022 [53] | India | 10-35 | 38 | LAmB (5mg/kg, twice weekly for three weeks) | Unknown | Undefined | Undefined |
| 5 | Singh OP et al. 2021 [17] | India | 12 – 64 | 26 | Miltefosine (100mg/day for 12 weeks) | 180 | Undefined | Undefined |
| 6 | Younis et al. 2021 [54]<br>(Trial registration: NCT02894008) | Sudan | 12–37 | 24 | Arm 1: sterile aqueous buffered solution containing ChAd63-KH at a concentration of 10 billion viral particles as a single dose in 1mL volume intramuscularly into the deltoid muscle. | 120 | Not stated | Not stated |
|  |  |  |  |  | Arm 2: A sterile aqueous buffered solution containing ChAd63-KH at a concentration of 75 billion viral particle/mL was administered as a single dose in 1mL volume intramuscularly into the deltoid muscle. |  |  |  |
| 7 | Khalil EA et al. 1996 [55] | Sudan | 9 – 32 | 9 | Terbinafine 250mg/day for 4 weeks | 28 | PKDL grade |  |
| 8 | Ganguly S et al. 2010 [56] | India | 12 – 65 | 21 | Arm 1: Sodium stibogluconate (20mg/kg/day intramuscular for 3 months)<br>Arm 2: Miltefosine (100mg/day per oral for 2 months) | 30 | Undefined | Remission of clinical features was a primary criterion of cure |
| 9 | Islam S et al. 2013 [57] | Bangladesh | 2 – 68 | 87 | Arm 1: Sodium stibogluconate (20mg/kg/day in intramuscular route for a total of 6 cycles of treatment, each cycle consisting of 20 days of treatment with an interval of 10 days in between two cycles)<br>Arm 2: LAmB (5mg/kg two times per week for 3 weeks) | 2190 | Undefined | Resolution of PKDL lesions was confirmed by physician examination |
| 10 | Neogy AB et al. 1988 [58] | India | 12 – 52 | 10 | Sodium antimony gluconate (45 injections at 10mg/kg body weight once/day administered on alternate days) | 180 | Undefined | Undefined |
| 11 | Mukherjee S et al. 2015 [59] | India | Unknown | 10 | Arm 1: Sodium antimony gluconate (20mg/kg/day for 4 months)<br>Arm 2: Miltefosine (100mg/day for 4 months) | 120 | Undefined | Undefined |
| 12 | Mukherjee S et al. 2019 [60] | India | Unknown | 20 | Arm 1: Sodium antimony gluconate (20mg/kg/day for 4 months)<br>Arm 2: Miltefosine (100mg/day for 4 months) | Unknown | Undefined | Undefined |
| 13 | Mukhopadhyay D et al. 2016 [24] | India | Unknown | 87 | Arm 1: Sodium antimony gluconate (20mg/kg/day for 4 months)<br>Arm 2: Miltefosine (100mg/day for 4 months) | 180 | Undefined | Undefined |
| 14 | den Boer et al. 2018 [13] | Bangladesh | Not stated | 280 | L-AmB (15 mg/kg in 5 doses of 3 mg/kg, biweekly dosing) | 365 | Not stated | Weighed percentage of improvement and descriptive categories at 12 months<br>Category 1: Complete resolution of nodular and papular lesions, and complete or almost complete re-pigmentation of macular lesions<br>Category 2: Complete resolution of nodular and papular lesions and major re-pigmentation of macular lesions; some macular lesions have resolved<br>Category 3: No or limited improvement of lesions, but no new lesions<br>Category 4: No or limited improvement of lesions with emergence of new lesions. |
| 16 | Bhandari V et al. 2012 [61] | India | Unknown | 8 | Miltefosine (50mg capsule, thrice daily for 60 days) | 365 | Undefined | Undefined |
| 17 | Neogy AB et al. 1990 [62] | India | Unknown | 10 | Sodium antimony gluconate (10mg/kg daily for 3 months) | 240 | Undefined | Undefined |
| 18 | El Hassan AM et al. 1992 [63] | Sudan | 2 – 30 | 19 | Sodium stibogluconate (20mg/kg daily for 30 days, i.v.) | 365 | Undefined | Undefined |
| 19 | Das ML et al. 2007 [64] | Nepal | Unknown | 25 | Sodium antimony gluconate (20mg/kg/day)* | 365 | Undefined | Undefined |
| 21 | Musa AM et al. 2002 [7] | Sudan | Unknown | 7 | Sodium stibogluconate (20mg/kg/day i.v. for three months) | Unknown | Undefined | Undefined |
| 22 | Katara GK et al. 2011 [65] | India | 19 – 52 | 25 | Miltefosine (150mg/day for 2 months) | 30 | Undefined | Undefined |
| 23 | Basher A et al. 2019 [66] | Bangladesh | 16 – 45 | 29 | Miltefosine (2.5mg/kg/day for 12 weeks. Patients ≥25kg received 100mg per day) | Unknown | Undefined | Undefined |
| 24 | Ganguly S et al. 2015 [67] | India | 5 – 42 | 26 | Sodium stibogluconate (20mg/kg, i.m., for 90-120 days) | 545 | Undefined | A cure case is defined as complete disappearance of skin lesion(s) after treatment, as reported by the patient and assessed by the clinician and also negative by PCR. |
| 26 | Hossain F et al. 2017 [68] | Bangladesh | Unknown | 40 | Miltefosine, 100mg/day, orally for 12 weeks for patients weighing >25 kg and 50mg orally per day for 12 weeks for patients weighing <25kg | Unknown | Undefined | PKDL cases after completing treatment and disappearance of any visible skin rash were considered cured |
| 27 | Zijlstra EE et al. 1993 [69] | Sudan | 3 – 40 | 2 | Sodium stibogluconate at 20mg/kg/day for 30 days | 365 | Undefined | Undefined |
| 28 | Modak D et al. 2010 [70] | India | 13 – 38 | 6 | Miltefosine capsule (50 mg) twice daily for 8 weeks | 365 | Undefined | Undefined |
| 29 | Ghosh P et al. 2018 [71]<br>(Trial registration: NCT02193022) | Bangladesh | Unknown | 83 | Miltefosine allometric dose | 365 | Undefined | Undefined |
| 30 | Khalil EA et al. 2002 [72] | Sudan | Unknown | 47 | Sodium stibogluconate for 1 to 2 months ** | Unknown | Undefined | Undefined |
| 31 | Ramesh et al. 2015 [73] | India | ≥8 | 86 | Arm 1:<br>Miltefosine (50 mg twice daily for 3 months and 2.5 mg/kg/day for 3 months in children.)<br>Arm 2:<br>Miltefosine (50 mg thrice daily for 2 months for adults.) | 540 | Treatment completed, clinical regression (total regression of papules/ nodules with no new lesion for polymorphic PKDL and lack of appearance of new lesions in macular PKDL), parasitological cure indicated by a negative smear and/or insignificant parasite burden by real time PCR at one-month post MIL therapy. | PKDL case with initial cure and no clinical signs (total regression of papules/ nodules, no new lesion and near total re-pigmentation of maculae) at 18 months after completion of therapy. |
| 32 | Das S et al. 2017 [74] | India | Unknown | 46 | Amphotericin B (1mg/kg body weight as alternate-day infusions for 40 days for 5 months with two 15-day breaks between the courses) | Unknown | Undefined | Cure was assessed by the disappearance of clinical features, i.e. the lesion and/or clearance of parasite in the lesion. |
| 33 | Mukherjee S et al. 2016 [75] | India | Unknown | 34 | Arm 1: Sodium antimony gluconate (20mg/kg/day for 4 months)<br>Arm 2: Miltefosine (100mg/day for 4 months) | 120 | Undefined | Undefined |
| 35 | Jaiswal P et al. 2018 [76] | India | 5 – 60 | 90 | Arm 1: LAmB (twice weekly at 5mg/kg body weight for three weeks)<br>Arm 2: Miltefosine (50mg/kg body weight for twelve weeks) | 181 | Not presented | Clinical cure and absence of dermal lesions |
| 36 | Zijlstra EE et al. 1994 [77] | Sudan | Unknown | 52 | Sodium stibogluconate (20mg/kg, daily intramuscular injections) | 365 | Undefined | Healing of lesions (Table 2) |
| 37 | Sundar et al. 2014 [78] | India | Not stated | 31 | Paromomycin 11mg/kg intramuscularly for 45 days daily. | 365 | Not stated | Cure was defined as complete disappearance of skin lesion(s) after treatment, as reported by the patient and assessed by the trained physician at 12-month follow-up. |
| 39 | Sundar et al. 2015 [79] | India | Not stated | 33 | Miltefosine was administered at a target dose of 2.5mg/kg/day for 12 weeks. Patients $\geq 25$ kg received 100mg per day: | 365 | Not stated | Cure was defined as complete disappearance of skin lesion(s) after treatment, as reported by the patient and assessed by the |
|  |  |  |  |  | one 50mg capsule in the morning and evening with meals. Patients <25 kg received 50mg/day: one 50mg capsule per day. The patients were treated as inpatients for the first 4 weeks and then continued as outpatients for the rest of the treatment period. |  |  | trained physician at 12-month follow-up. |
| 40 | Ghosh et al. 2015 [80] | India | 10–60 | 27 | Miltefosine (50 mg in a daily single dose of 100 mg/day (body weight ≥ 25 Kg) or 50 mg/day (body weight < 25 Kg) or 2.5 g/kg/day (aged 2-11 years). All patients were treated for 16 weeks unless treatment-emergent side effects limited further therapy; in case of polymorphic PKDL, therapy was terminated earlier, if there was complete resolution of papulo-plaques/nodules. | 2373 | Not stated | Undefined |
| 41 | Garg VK et al. 2001 [81] | Nepal | 4 – 62 | 22 | Sodium antimony gluconate (intramuscular, 20mg/kg/day for a period ranging from 30 to 72 days depending upon the clinical response) | 72 | Undefined | Undefined |
| 42 | Ramesh V et al. 1993 [82] | India | 10 – 50 | 18 | Sodium antimony gluconate (daily intramuscular, 20mg/kg body weight, total dose not exceeding 1g/day) | 1800 | Undefined | Undefined |
| 45 | Ganguly S et al. 2008 [83] | India | 6 – 47 | 18 | Sodium antimony gluconate (20mg/kg/day for 3 months) | 30 | Undefined | Assessment of cure was based on remission of clinical features |
| 46 | Verma S et al. 2013 [84] | India | 9 – 50 | 74 | Arm 1: Sodium antimony gluconate (20mg/kg/day) for 4 months<br>Arm 2: Miltefosine (150mg/day) for 2 months | 365 | Undefined | Detection of parasites in the slit aspirate |
| 47 | Moulik S et al. 2018 [85] | India | Unknown | 182 | Arm 1: Miltefosine (50mg orally twice daily for 12 weeks)<br>Arm 2: LAmB (5mg/kg twice weeklyfor3weeks) | 180 | Undefined | The clinical outcome was assessed by a dermatologist and they were considered cured based on total regression of papules/nodules, no new lesion(s), and considerable regression of macular lesions |
| 48 | Ramesh et al. 2020 [86] | India | 9–60 | 32 | Arm 1:<br>Miltefosine at a dose of 100mg/day, except for 3 children between the age of 9 and 12 years weighing <25 kg, who were administered at a dose of 50mg/day after meals for a period of 90 days<br>Arm 2:<br>Three i.v. injections of L-AmB (5 mg/kg body weight) given on days 1, 8, and 15. + Miltefosine capsules were administered after the first dose of L-AmB and continued daily until the end of 45 days | 540 | Not stated | Defined on the basis of clinical examination with total regression of papules/nodules and near total re-pigmentation of maculae at 18 months follow up. Nil/negligible parasite load by real time PCR. |
| 49 | Ramesh et al. 2011 [87] | India | 17–60 | 26 | Miltefosine (50 mg thrice daily were given after food for 60 days; treatment was extended by up to a maximum of 30 days if a responder showed incomplete cure | 365 | The presence of parasites was evaluated from lesioned sites at 1-month post-treatment using qPCR | Subsidence of indurated lesions leaving normal or wrinkled skin, signs of re-pigmentation and histopathological absence of disease constituted cure. |
| 52 | Singh RP et al. 2012 [88] | India | 10 – 34 | 6 | Arm 1: Miltefosine 100mg/day and 50mg/day for 84 days<br>Arm 2: Amphotericin B (1mg/day for 30 days, every alternate day) | Unknown | Undefined | Regression of lesion (Table 1 of the manuscript) |
| 53 | Chowdhury S et al. 1991 [89] | Bangladesh | Unknown | 45 | Sodium antimony gluconate (20mg/kg for six courses of 20 days with 10 days in between each two courses) | 365 | Undefined | All treated VL and PKDL cases were then assessed for response to treatment during follow-up examinations conducted three and 12 months after treatment, in which the post treatment parasitological results, clinical and haematological improvements were all evaluated. |
| 54 | Elhassan AM et al. 1992 [90] | Sudan | Unknown | 18 | Sodium stibogluconate (20mg/kg intravenous daily for 30 days) | Unknown | Undefined | Undefined |
| 55 | Jensen AT et al. 1999 [91] | Sudan | 8 – 47 | 26 | Sodium stibogluconate (daily dose of 20mg/kg for 2 months) | 60 | Undefined | Undefined |
| 56 | Ismail A et al. 1999 [92] | Sudan | 8 – 42 | 19 | Sodium stibogluconate (20 mg/kg/day intravenous for 2 months) | 180 | Undefined | healing of lesions |
| 60 | 60 | Thakur et al. 1990[93] | India | 4-60 | 53 | Sodium stibogluconate, at the dose of 20 mg/kg/body weight/day/ i.m. with a maximum of 8.5 mL) for 120 days (or more, if necessary) | 365 | Not stated |
| 71 | Sengupta et al. 2021[94] | India | Unknown | 101 | Miltefosine for 12 weeks (precise description of regimen is unknown) | 180 | Unknown | Unknown |

\*Duration of treatment was unknown.
\*\*Dose of treatment was unknown.
i.v. = intravenous; i.m. = intramuscular; LAmB = Liposomal amphotericin B

**Table 4:** Summary of the drug regimens adopted in PKDL studies.

| Drug regimen | Number of studies | Number of arms | Number of patients | Percentage |
| --- | --- | --- | --- | --- |
| Miltefosine | 25 | 27 | 636 | 33.9% |
| Liposomal amphotericin-B (L-AmB) | 7 | 7 | 508 | 27.1% |
| Sodium stibogluconate (SSG) | 15 | 17 | 268 | 14.3% |
| Sodium antimony gluconate (SAG) | 13 | 13 | 186 | 9.9% |
| Amphotericin-B deoxycholate (AMBd) | 2 | 2 | 57 | 3.0% |
| Amphotericin B (unclear formulation) | 2 | 3 | 53 | 2.8% |
| Miltefosine + Paromomycin | 1 | 2 | 45 | 2.4% |
| Paromomycin | 1 | 1 | 31 | 1.7% |
| ChAd63-KH vaccine | 1 | 2 | 24 | 1.3% |
| L-AmB + Miltefosine | 1 | 1 | 16 | 0.9% |
| Alum/ALM + BCG + SSG | 1 | 1 | 15 | 0.8% |
| SSG + placebo vaccine | 1 | 1 | 15 | 0.8% |
| Terbinafine + Itraconazole | 1 | 1 | 9 | 0.5% |
| SAG + Ketoconazole | 1 | 1 | 5 | 0.3% |

BCG = Bacillus Calmette–Guérin vaccine; Alum/ALM= Alum Precipitated Autoclaved Leishmania major (Alum-ALM) vaccine

### Reporting of safety data (n=56 studies)

Adverse treatment outcomes were reported in all 8 RCTs. The Common Terminology Criteria for Adverse Events (CTC-AE) of the National Cancer Institute (NCI) of the National Institutes of Health (NIH), USA, was used in 3 RCTs and the AE methodology, criteria and terminology was not clear in 5 RCTs. Of the 48 non-randomised studies, CTC-AE was used in 5 studies and not reported in the remaining 43 studies.

## Discussion

This review identified 56 studies (2,486 patients) published in the past 50 years describing the efficacy of drug regimens adopted for treatment of PKDL. Of these, only 8 studies representing 15% of the enrolled PKDL patients were from randomised clinical trials. Despite the fact that PKDL is a potential reservoir of infection [16, 23] with more than 700 to 2,322 cases reported annually (2014 – 2021)[23], identification of only eight RCTs studies suggests a relatively limited evidence base supporting treatment recommendations. This review also identified that there were a limited number of studies among children (in studies that reported patients’ age-range) with an evident male preponderance (male: female ∼ 2:1) among enrolled patients; this is consistent with published literature on PKDL and VL [24-26]. None of the 56 studies included in this review enrolled pregnant or lactating women. In general, PKDL in pregnancy has been described only in sporadic case reports in the literature [27]. Similar observations were found for the treatment of VL in pregnancy [28]. Finally, none of the studies identified enrolled patients with HIV or other comorbidities. Although these comorbidities are commonly reported in visceral leishmaniasis [29, 30], these conditions have been less pronounced with PKDL, and only sporadic cases have been reported across India, Africa, Europe, and South America [31-33]. Overall, these observations regarding the patient spectrum show substantial knowledge gaps in treatment efficacy in these specific subpopulations.

Of the 48 observational studies, three (125 patients) studies did not require any confirmatory tests for the PKDL diagnosis. All the RCTs used one or more laboratory tests for the PKDL diagnosis. Serological tests (antibody detection) were used in six studies involving 471 patients, although such tests are not deemed adequately reliable in this context as there can be substantial false-positive test results due to past VL infection and antibody cross-reactivity [34]. Serological tests started to be adopted for diagnosis after the 1990s and have continued to be implemented in PKDL studies, particularly, in combination with microscopy and clinical criterion. The rK39 rapid immunochromatographic test was the most common serological test used while enzyme-linked immunosorbent assay and the direct agglutination test were also reported. Microscopic demonstration of amastigotes in skin biopsies (slit or snip skin smears) was adopted throughout the time-period included in this review. In studies published after 2010, PCR method has been used, particularly in combination with microscopy. However, the choice of PCR types (nested, quantitative, etc.) and target genes (ITS-1, K-39) varied substantially across studies.

Following enrolment of patients in 82 arms across 56 studies, 13 different drugs were used for treatment with more than 85% of the patients treated with three major drugs: liposomal amphotericin B, miltefosine, and pentavalent antimony. However, even for these major drugs, there was a wide variation in daily mg/kg dose and overall treatment duration. The adoption of drug regimens was also highly regional. Miltefosine and L-AmB were adopted in the Indian subcontinent, whereas pentavalent antimony was the major drug used in Sudan. Following the completion of the treatment regimen, there was also variation in the time-point of assessment of cured proportion, including differences in criterion used for defining cure. In general, the most common definition of initial cure was parasitological clearance after treatment completion with or without clinical resolution of lesions at the planned end of the completion of the treatment regimen. Similarly, complete resolution of lesions with parasitological clearance at the time of final assessment (actual time-point of evaluation varied) was the most common definition of final cure.

Overall, this review has characterised the variation in different aspects of study design, conduct, analysis, and reporting. For example, the outcome definitions adopted for efficacy assessments were presented in only a third of the studies, the age-range of the patients included was not reported in 43% of the studies while the diagnostic method for disease confirmation was unclear in 11% of the studies. This heterogeneity acted as a limitation to perform a direct comparison of results across the studies using the published aggregated data. Similarly, criteria used for gradation of the severity of safety outcomes were not reported in the majority of the studies included; an observation consistent with the VL literature [35]. Taken together, this warrants harmonisation of the definitions and protocols adopted across future studies for facilitating robust reporting of the efficacy and safety studies of PKDL. Besides, a scope for harmonisation of these aspects in the existing data can be exercised through IPD data platform.

Similarly, the median sample size recruited across 8 RCTs and 48 non-randomised studies were 33 (range 10 – 108) and 26 (range 2 – 289), respectively. This suggests that existing practice on the patient treatment of PKDL relies on evidence from relatively small studies. Such small study size including wide variations in the regimens adopted across the studies limits the possibility of robust assessment of efficacy and to identify determinants of poor treatment outcomes in any single study. Individual patient data (IPD) meta-analyses are a proven methodological approach to address some of the limitations of classical aggregated meta-analyses [36-38]. A harmonised repository of raw data from the studies identified in this review can facilitate IPD meta-analyses to generate new evidence regarding the efficacy of existing medicines in sub-groups of patient population. Understanding the existing volume of data was a crucial first step towards determining the feasibility of establishing an IPD data repository for PKDL. First, over half of the studies on PKDL were published in the last decade. The data from these recent studies are more likely to be retrievable than the older studies. Second, the identified volume of existing data on miltefosine and liposomal amphotericin B in the trials (>500 patients in total) (these new regimens were mostly tested in studies after 2000) suggests that creating an IPD platform for these two important drug regimens would be valuable. For studies investigating treatment regimens with fewer numbers of enrolled patients, such as paromomycin, assembling the raw IPD across multiple studies could help to provide further information on these regimens. Finally, a new large study has been published since the completion of this review [39], which can be incorporated in the future update of this systematic review.

The Infectious Diseases Data Observatory (IDDO) has an established experience in the development of an IPD repository for <u>VL</u> and other infectious diseases, including but not limited to malaria [<u>WWARN</u>], <u>Ebola Virus Disease and COVID-19</u>. These IDDO repositories have facilitated large scale IPD meta-analyses exploring the determinants of therapeutic efficacies [40-42]. To maximise interoperability and reusability of data, IDDO has adopted the Clinical Data Interchange Standards Consortium (CDISC) compliant data standards [43]. As an important step towards the development of a PKDL data platform, the <u>IDDO VL dataplatform</u> already hosts a critical mass of data from efficacy trials in VL conducted over the past 20 years and hence has the technical and infrastructural framework to support the development of a PKDL data platform. Based on the identified volume of data with a substantial number of studies being relatively recent, and initial sharing of datasets from some key PKDL studies to the IDDO VL data platform suggests that establishment of a PKDL data platform is feasible. Such a platform can facilitate collaborative research to answer questions of public health relevance and maximise the usefulness of existing data and promote data re-use. The unique resource that would be created by a harmonised database of IPD from the identified studies would allow us to answer issues of public health concern that could not be answered through standalone trials or aggregated published data.

## Conclusions

An IPD platform would have enough data on the three main medications (miltefosine, L-AmB, and pentavalent antimony) if the number of patients from nonrandomised studies were pooled, despite the fact that this review found insufficient patient volume across randomised clinical trials. With the help of relevant stakeholders, the global PKDL community, and sufficient financing, a PKDL IPD platform can be realised. This will allow a nuanced exploration of the safety and efficacy of PKDL drugs which can potentially guide future clinical practices.

## Declarations

### Authors’ contributions

Conceptualisation: BJM, KP, PD, PJG, SSP

Literature Search: EH

Data Curation: AM, RK, RC, SR, SSP

Formal Analysis: RK, PD, SSP

Funding Acquisition: PJG

Investigation: RK, AM, PD, RC, SR, SSP

Methodology: BJM, PD, SR, SSP

Project Administration: CN, SSP

Resources: KP, PJG

Software: AM, CN, PD, RC, RK, SR

Supervision: KP, NAS, PD, PJG, SSP

Validation: RK, AM

Visualisation: PD

Writing – Original Draft Preparation: RK, PD, SSP

Writing – Review & Editing: All

### Availability of data and material

The data from this systematic review are available in the supplemental files.

### Ethics approval and consent to participate

Not applicable

### Consent for publication

Not applicable

### Financial Disclosure Statement

The review was funded by a biomedical resource grant from BMGF to the Infectious Diseases Data Observatory (Recipient: PJG; Grant Number: INV-004713). The funders had no role in the design and analysis of the research or the decision to publish the work.

### Competing interests

None

## Data Availability

All data produced in the present work are contained in the manuscript supplement files

## Notes

### Competing Interest Statement

The authors have declared no competing interest.

### Funding Statement

The review was funded by a biomedical resource grant from the BMGF to the Infectious Diseases Data Observatory (Recipient: PJG; Grant Number: INV-004713).

