## Supplemental File 2 for "Post-kala-azar dermal leishmaniasis (PKDL) drug efficacy study landscape: a systematic scoping review of clinical trials and observational studies to assess the feasibility of establishing an individual participant-level data (IPD) platform"

**Supplemental file 2: Search strategies**

Post-kala-azar dermal leishmaniasis (PKDL) drug efficacy study landscape: a systematic scoping review of clinical trials and observational studies to assess the feasibility of establishing an individual participant-level data (IPD) platform

Sauman Singh-Phulgenda^1,2*§^, Rishikesh Kumar^3§^, Prabin Dahal^1,2^, Abdalla Munir^4^, Sumayyah Rashan^1,2^, Rutuja Chhajed^1,2^, Caitlin Naylor^1,2^, Brittany J. Maguire^1,2^, Niyamat Ali Siddiqui^3^, Eli Harriss^5^, Manju Rahi^6^, Fabiana Alves^7^, Shyam Sundar^8^, Kasia Stepniewska^1,2^, Ahmed Musa^4^, Philippe J Guerin^1,2^, Krishna Pandey^3*^

### Systematic search of clinical literature

Eli Harriss (Bodleian Health Care Libraries, University of Oxford, ORCID: 0000-0003-4635-8959) developed the search strategy and syntax, advised on the databases to use, ran the searches on 02/06/2020 and ran the update searches in full on 03/03/2023, deduplicated the records, and reviewed the pre-submission manuscript.

#### Search Results

|  | Search results 02/06/2020 | Search results 05/10/2021 | Search results 03/03/2023 |
| --- | --- | --- | --- |
| PubMed | 539 | 595 | 627 |
| Ovid Embase | 608 | 662 | 715 |
| Ovid Global Health | 479 | 524 | 559 |
| Scopus | 589 | 648 | 677 |
| Web of Science Core Collection | 492 | 550 | 575 |
| Cochrane Database and CENTRAL | 32 | 35 | 36 |
| PASCAL | 123 | 123 | 123 |
| WHO Global Index Medicus | 60 | 63 | 63 |
| clinicaltrials.gov | 17 | 17 | 18 |
| WHO ICTRP |  |  | 20 |
| Indian Clinical Trials Registry |  |  | 5 |
| Total | 2939 | 3217 | 3418 |
| Total after deduplication | 848 | 923 | 977 |
| Unique to databases since previous search date |  | 94 | 67 |

#### Search Strategies

**PubMed**

((Post-Kala-azar Dermal) OR pkdl) OR (PostKalaazar) ALL FIELDS

**Database: Embase 1974 to present**

1. "Post-Kala-azar Dermal".mp. [mp=title, abstract, heading word, drug trade name, original title, device manufacturer, drug manufacturer, device trade name, keyword, floating subheading word, candidate term word]

2. PKDL.mp.

3. 1 or 2

**Database: Global Health 1973 to 2020 Week 21**

1. "Post-Kala-azar Dermal".mp. [mp=title, abstract, heading word, drug trade name, original title, device manufacturer, drug manufacturer, device trade name, keyword, floating subheading word, candidate term word]

2. PKDL.mp.

3. 1 or 2

**Scopus:**

TITLE-ABS-KEY ( "Post-Kala-azar Dermal"  OR  pkdl )

**Web of Science Core Collection**

TOPIC: ("Post-Kala-azar Dermal" OR pkdl)

**Cochrane Database of Systematic Reviews**

**Issue 3 of 12, March 2023**

**Cochrane Central Register of Controlled Trials**

**Issue 2 of 12, February 2023**

#1	"Post-Kala-azar Dermal" OR pkdl	36

4 Cochrane Database of Systematic Reviews

32 Cochrane Central Register of Controlled Trials

**Pascal and Francis Bibliographic Databases**

<https://pascal-francis.inist.fr/vibad/index.php?action=search&lang=en&terms=%22Post-Kala-azar+Dermal%22++OR++pkdl>+

"Post-Kala-azar Dermal" OR pkdl

**WHO Global Index Medicus** [**https://pesquisa.bvsalud.org/gim/?lang=en**](https://pesquisa.bvsalud.org/gim/?lang=en)

Title, abstract, subject: "Post-Kala-azar Dermal" OR pkdl

<https://pesquisa.bvsalud.org/gim/?lang=en&_charset_=utf-8&index=&filter%5Bdb%5D%5B%5D=&q=%22Post-Kala-azar+Dermal%22++OR++pkdl>

**Clinicaltrials.gov – condition or disease**

Post-Kala-azar Dermal

PKDL

**WHO International Clinical Trials Registry Platform** <https://trialsearch.who.int/AdvSearch.aspx> – searched 03/03/2023

(Title OR Condition): Post-Kala-azar Dermal OR pkdl

Recruitment status is: ALL

2 studies registered from 2021-2023; 20 in total.

**Indian Clinical Trial Registry** <http://ctri.nic.in/Clinicaltrials/pubview.php>

Searched 03/03/2023

Health Condition/ Problem Studied: Post-Kala-azar Dermal = 0 results

Health Condition/ Problem Studied: PKDL = 5 results
